# Neurochemical fingerprinting of the Alzheimer’s disease continuum using 7 T short-echo-time brain single voxel ^1^H-MRS

**DOI:** 10.64898/2026.10.16.26365733

**Authors:** Andrea Dell’Orco, Laura Göschel, Layla Tabea Riemann, Semiha Aydin, Agnes Flöel, Anna Tietze, Michael Scheel, Ariane Fillmer

## Abstract

**Introduction:** This study aimed to characterize the neurochemical profile of Alzheimer’s disease (AD) continuum stages using proton magnetic resonance spectroscopy (^1^H-MRS) data for brain metabolites and macromolecules. We used Orthogonal Projection to Latent Structures Discriminant Analysis (OPLS-DA) to classify diagnostic groups and assess feature importance.

**Methods:** Our dataset consisted of 7 T short-echo time ^1^H-MRS spectra of the posterior cingulate cortex/precuneus of individuals diagnosed with AD (n = 27) or Mild Cognitive Impairment (MCI, n = 27) and Healthy Controls (HC, n = 31). Classification used 100-round Monte Carlo cross-validation based on metabolite concentrations and macromolecular peak intensities. Variable importance in projection (VIP) was assessed across classification models to identify the most relevant neurochemical features. The resulting metabolite rankings were compared with previous literature to evaluate their biological plausibility.

**Results:** Classification was strongest for AD versus HC (AUC=0.88) and lower for comparisons involving MCI. Metabolites contributing most strongly to classification included myo-inositol, ascorbate, and choline-containing compounds, consistent with previous studies on AD-related neurochemical alterations. In addition, specific macromolecular components were identified as relevant features, suggesting a potential contribution of MM signals to the characterization of AD-related neurochemistry.

**Discussion:** These findings support the use of short-echo-time ^1^H-MRS as a complementary tool for characterizing neurochemical changes in the AD continuum, with potential applications in disease characterization and monitoring, warranting validation in larger cohorts.

## Introduction

Alzheimer’s disease (AD) is a progressive and fatal neurodegenerative disorder that leads to dementia [1]. The disease advances gradually, spanning a continuum of clinical stages from normal functioning to AD. Mild cognitive impairment (MCI) is considered a prodromal stage with a high risk of progressing to AD [2]. Early and reliable diagnosis and accurate monitoring are crucial for successful interventions, e.g., recently approved treatments target amyloid beta plaques that accumulate in the pre-clinical phase of the disease [3].

AD pathology can be assessed using molecular biomarkers such as amyloid/tau PET and CSF-based measures, but these approaches may be limited by invasiveness, cost, or accessibility. Additional non-invasive biomarkers are therefore needed to support AD diagnosis, treatment selection, and disease monitoring. ^1^H-MRS is a non-invasive technique that provides insights into the brain’s neurochemistry. Previous studies have reported AD-related alterations in metabolites such as myo-inositol (mIns), total N-acetylaspartate (tNAA), total choline (tCho), and total creatine (tCr)[4]. However, there is currently no broad consensus regarding the expected changes in the concentrations of these metabolites in AD[4,5].

Short-TE ^1^H-MRS can reveal not only metabolites but also signals from cytosolic proteins, commonly referred to as macromolecules (MM). Some evidence suggests a possible role of MM signals as a biomarker [6]. To the best of our knowledge, the macromolecular profile across the AD continuum has not yet been systematically characterized.

Because metabolites are embedded in interconnected biochemical networks, multivariate approaches that jointly model multiple features may provide improved sensitivity and biological interpretability. Orthogonal projection to latent structures discriminant analysis (OPLS-DA[7]) enables classification while retaining interpretability through the measure of the VIP.

Previous works have demonstrated the utility of multivariate classifiers, for example, Marjańska et al.[8] utilized MRS data from the precuneus and occipital cortex, using Random Forest as a classifier, achieving sensitivity and specificity of 88% and 97%, respectively, in the AD vs healthy controls (HC) classification with Ins and ascorbate as main predictors. Additionally, Munteanu et al.[9] employed several classifiers and a combination of variables, achieving a maximum AUROC of 0.90 for the classification of AD vs HC using a mix of metabolite concentrations, metabolite ratios, and brain tissue volumes. Westman et al. (2010) investigated metabolite concentration ratios measured in the hippocampus of AD and HC involving Ins, tCho, and NAA, using Orthogonal Projection to Latent Structures Discriminant Analysis (OPLS-DA, Trygg & Wold, 2002). They achieved 76% and 83% sensitivity and specificity, respectively, for the classification of AD vs HC. OPLS-DA has also been effectively used for AD classification based on ^1^H-NMR concentrations of cerebrospinal fluid (CSF) metabolites [11].

The aim of this study was to characterize neurochemical alterations across the AD continuum using short-echo-time 7 T ^1^H-MRS data from the posterior cingulate cortex (PCC), including both metabolite concentrations and MM peak intensities. As an exploratory secondary aim, we evaluated whether these features, together with age, could distinguish diagnostic groups using OPLS-DA. We hypothesized that neurochemical profiles would show the strongest separation between AD and HC, whereas comparisons involving MCI would show greater overlap due to the intermediate and heterogeneous nature of MCI.

To our knowledge, this study is the first to model and analyze individual MM components in AD and to incorporate them into a multivariate classification model. Additionally, this study contributes to understanding the potential role of ^1^H-MRS data in early detection and monitoring of AD.

## Methods

### Participants

Our dataset originated from the projects NeuroMET (2016) and NeuroMET2 (2019), which focused on improving diagnosis and monitoring of neurodegenerative diseases using standardized methodologies. The ethics committee at Charité University Hospital approved the study (EA1/197/16 and EA2/121/19), and all procedures adhered to the principles outlined in the Declaration of Helsinki.

Diagnoses were assigned based on neuropsychological assessments. Exclusion criteria comprised: significant or unresolved medical, neurological, or psychological issues that might impact cognitive function, as well as those with past records of substance abuse or eating disorders. All participants provided informed consent before participating in the research. The present analysis was restricted to HC, MCI, and AD and the final dataset consisted of a total of 85 spectra: 31 from HC, 27 from MCI, and 27 from AD patients. Participants’ demographics are reported in Table 1.

**Table 1:** Participant demographics, Tissue fractions and spectral quality metrics. GM: gray matter, WM: white matter, CSF: cerebrospinal fluid, FQN fit quality number, FWHM: full width half maximum NAA peak. 1: one way ANOVA, 2: chi squared test. ^*^: p<0. 05, ^****^: p<0. 0001, n. s. non significant, based on non-adjusted p-values

**DEMOGRAPHICS**
|  | HC | MCI | AD | Total | Test Statistic | Significance |
| --- | --- | --- | --- | --- | --- | --- |
| **N** | 31 | 27 | 27 | 85 | 3 | n.s. |
| **AGE** | 71.8 ± 7.7 | 70.9 ± 5.2 | 74.1 ± 6.2 | 72.73 ± 7.10 | 3.88<sup>1</sup> | *<sup>1</sup> |
| **PERCENT FEMALE** | 48.5 | 26.0 | 51.8 | 42.5 | 6<sup>2</sup> | n.s.<sup>2</sup> |
| **GM FRACTION** | 0.67 ± 0.01 | 0.68 ± 0.01 | 0.60 ± 0.01 | 0.65 ± 0.01 | 16.46 | ****<sup>1</sup> |
| **WM FRACTION** | 0.25 ± 0.01 | 0.23 ± 0.01 | 0.24 ± 0.01 | 0.24 ± 0.01 | 0.63 | n.s.<sup>1</sup> |
| **CSF FRACTION** | 0.09 ± 0.01 | 0.09 ± 0.01 | 0.16 ± 0.01 | 0.11 ± 0.01 | 13.38 | ****<sup>1</sup> |
| **FQN** | 16.06 ± 12.24 | 13.88 ± 8.37 | 11.78 ± 7.61 | 14.06 ± 9.88 | 1.41 | n.s.<sup>1</sup> |
| **FWHM (×10-2) PPM** | 5.18 ± 1.12 | 4.86 ± 0.62 | 5.61 ± 1.02 | 5.19 ± 0.99 | 2.50 | n.s.<sup>1</sup> |
| **WATER FWHM** | 12.57 ± 3.75 | 14.13 ± 5.08 | 10.55 ± 3.72 | 12.62 ± 4.09 | 4.91 | *<sup>1</sup> |
| **SNR** | 50.45 ± 7.89 | 50.88 ± 7.91 | 48.19 ± 9.21 | 50.62 ± 8.90 | 0.80 | n.s.<sup>1</sup> |

### Acquisition of MRI and MRS data

MRI and MRS data were acquired on a 7 T whole-body MAGNETOM MRI scanner (Siemens Healthineers, Erlangen, Germany) equipped with a 1TX/32RX head coil (NOVA Medical, Wilmington, USA).

For high-resolution anatomical imaging, we employed a three-dimensional T1-weighted (T1w) Magnetization-Prepared 2 Rapid Acquisition Gradient Echoes (MP2RAGE) sequence [12] with a denoised reconstruction [13].

The 20 × 20 × 20 mm^3^ volume-of-interest (VOI) for MRS was positioned in the PCC region. Before acquiring single-voxel spectra, localized RF calibration and second-order B_0_ shimming were performed. We utilized the spin echo full intensity acquired localized (SPECIAL) sequence [14,15] with interleaved outer volume suppression combined with VAPOR water suppression [16].

The MRI and MRS acquisition parameters are reported in Supporting Materials Table S1.

### MRS processing and quantification

MRS preprocessing and quantification followed our previously described PRaMM workflow[17]. Briefly, spectra were coil-combined, frequency-aligned, averaged[18], and quantified with LCModel[19] using a simulated metabolite basis set together with the PRaMM macromolecular model. Tissue fractions were obtained from MP2RAGE segmentation and used for partial-volume correction. Metabolite concentrations were water-scaled, relaxation-corrected to achieve the absolute concentrations in mM, whereas MM signals were water-scaled and partial-volume corrected without relaxation correction. Spectral quality was assessed by visual inspection, CRLBs, and fit quality number; spectra with major artifacts or lipid contamination were excluded. Full preprocessing, basis-set composition, and quantification details are provided in the Supporting Information.

### Spectral quality assessment and inferential statistics

To assess whether differences in spectral quality between groups could bias the statistical analysis of the neurochemical differences, the analyses were organized as follows: first, spectral quality metrics were compared across diagnostic groups to assess potential diagnosis-related bias in data quality. Water linewidth and FQN were tested using one-way ANOVA, whereas metabolite CRLBs were tested using Kruskal–Wallis tests due to violation of homoscedasticity assumptions. Second, exploratory group differences in metabolite concentrations and MM peak intensities were assessed using ANCOVA with age and sex as covariates, followed by post-hoc pairwise testing where appropriate. P-values from the exploratory concentration analyses were adjusted using FDR correction, while post-hoc comparisons were adjusted using the Holm method. Third, diagnostic classification was evaluated using repeated MCCV-based OPLS-DA, with performance summarized by F1 score, sensitivity, specificity, accuracy, AUC, Q^2^, R^2^, and VIP-derived feature importance.

### Classification Models and Metrics

Statistical analysis was performed with R and its libraries ropls [20], tidyverse [21], rstatix [22], hrbrthemes [23], pROC [24], and VIM [25].

To perform multivariate classification, we employed OPLS-DA, a method widely used in metabolomics and other omics data analyses [26]. OPLS-DA extends partial least squares discriminant analysis by incorporating orthogonal signal filtering, which separates class-related variation from orthogonal sources of variability, thereby improving interpretability [7]. The model provides variable VIP, which quantify the contribution of each feature to class separation. Furthermore, OPLS-DA is robust to noisy data and performs well in settings where the number of variables exceeds the number of observations, making it well suited for ^1^H-MRS data [26].

Because OPLS-DA requires a complete feature matrix, we applied a missing-data strategy to avoid excluding entire participants or features because of individual missing values. Metabolites or MM with more than 10% of missing values were excluded from the classification model, while for metabolites and MM with few missing values, we applied k-Nearest Neighbor (kNN) based imputation, setting k=5 and using only participants with the same diagnosis for each imputation. Age was considered in the kNN model as well. Only data used for OPLS-DA were imputed; data used for other statistical analyses were not.

To classify individual diagnoses based on the MRS data, we trained OPLS-DA models and assessed the importance of different features in discriminating between diagnoses. We incorporated both metabolites and macromolecules as features with age, making this a novel exploration of the MM background in the progression of AD. Age was included as the only non-MRS feature because diagnostic groups differed in age and age-related changes in metabolite levels could otherwise be incorrectly attributed to disease-related metabolic differences.

To reduce the number of model features and minimize uncertainty arising from potential signal misassignment, overlapping MM peaks were combined by summing their intensities. This was applied to M1.22 and M1.43, M2.05 and M2.27, and M3.59, M3.71, and M3.90.

A Monte Carlo Cross Validation (MCCV, each 100 rounds) was performed to ensure the model’s generalizability and robustness for the three following diagnostic pairs: AD vs HC, MCI vs HC, and AD vs MCI. The only non-^1^H-MRS variable included in the model was age.

A preliminary analysis was performed to determine the minimum number of orthogonal components required to improve predictive performance[27]. Each MCCV iteration was trained with class-balanced training sets by randomly selecting approximately 85% of participants from the smaller group and an equal number of participants from the larger group.

Classification performance metrics, such as specificity, sensitivity, AUC, F1 score and accuracy, were employed to assess the performance, considering their distribution throughout the MCCV process. The OPLS-DA model fit and predictive ability were evaluated using R^2^X, R^2^Y, and Q^2^. Furthermore, we computed averaged VIP (aVIP) for each metabolite by averaging individual VIP across MCCV rounds and using a threshold of one.

Additionally, we examined subject-wise misclassification frequency. Consistent errors may indicate neurochemical outliers or diagnostic discrepancies, whereas random errors may reflect suboptimal filtering or model underfitting.

## Results

### MRS Preprocessing and quantification

Acquisition parameters are summarized in Supporting Information Table S1. Visual inspection of the preprocessed spectra revealed one case of lipid contamination and one case of major artifacts, resulting in the exclusion of those spectra and their replacement with the available follow-up measurement of the same patient. Excluded spectra are reported in Supporting Information, Section S2.

Exemplary MRS voxel positioning and averaged spectra for each diagnosis are shown in Figure 1, respectively. AD participants are significantly older than MCI or HC. Compared with HC and MCI, the AD group exhibited a significantly lower gray matter (GM) fraction and a higher cerebrospinal fluid (CSF) fraction, consistent with greater atrophy. The average LCModel-derived spectral FWHM was 0.05 ppm (~15 Hz). No significant diagnosis effects were observed for FQN, LCModel-derived FWHM, SNR, or metabolite/MM CRLBs. In contrast, water FWHM showed a significant overall diagnosis effect (F = 4.91, p = 0.01).

**Figure 1:**
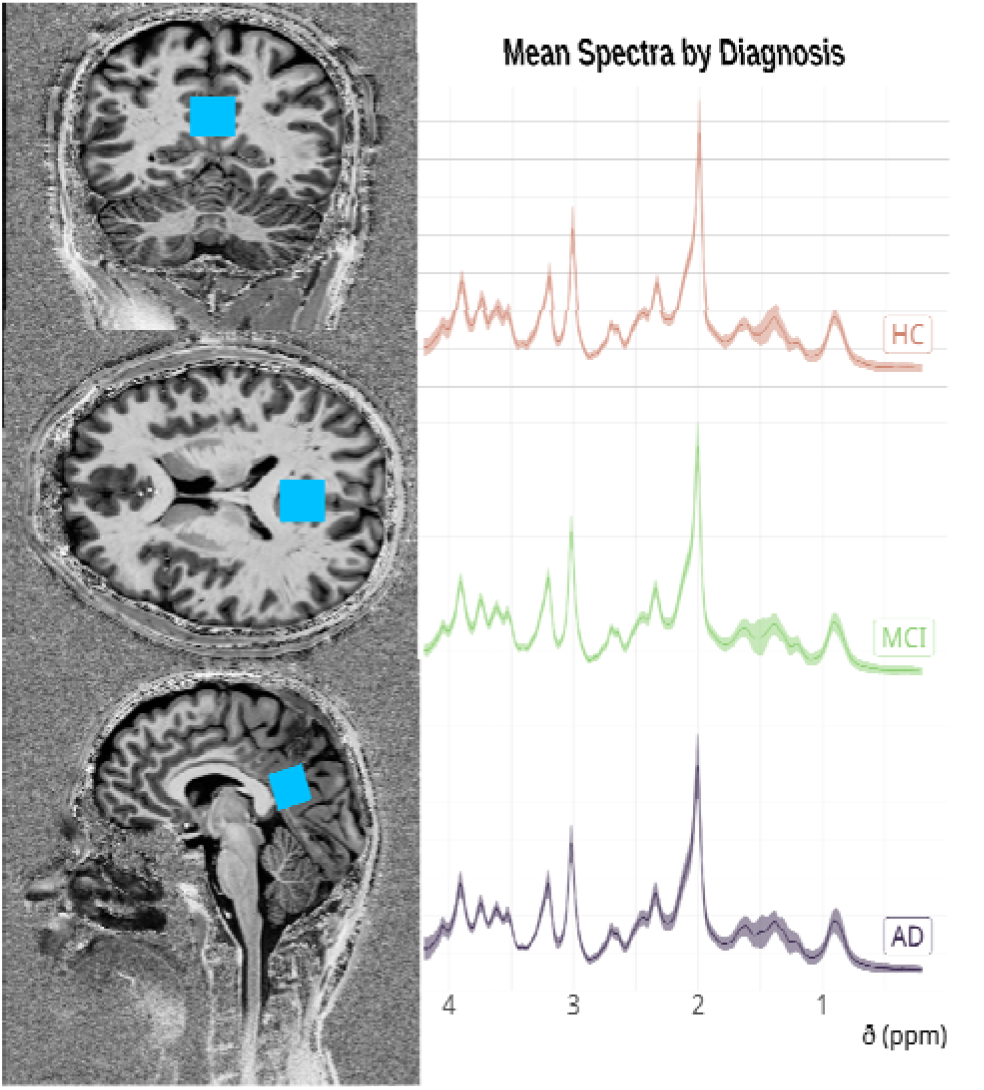
Voxel positioning and averaged PCC spectra for each diagnosis, shadowed areas represent the standard deviations. It can be seen, that each diagnosis has a high variance in chemical shifts between 1. 2 and 1. 8 ppm. Moreover, the ratio between the main peaks ofthe tCr and tCho at 3. 00 and 3. 20 ppm seems slightly altered in AD with respect to MCI or HC. The main peaks ofmIns at 3. 6 ppm appear higher relative to the closest peaks in MCI and AD as well.

Metabolites and MM average levels are shown in Figure 2, and the corresponding group-wise values together with ANCOVA results are reported in Supporting Information Table S3.

**Figure 2:**
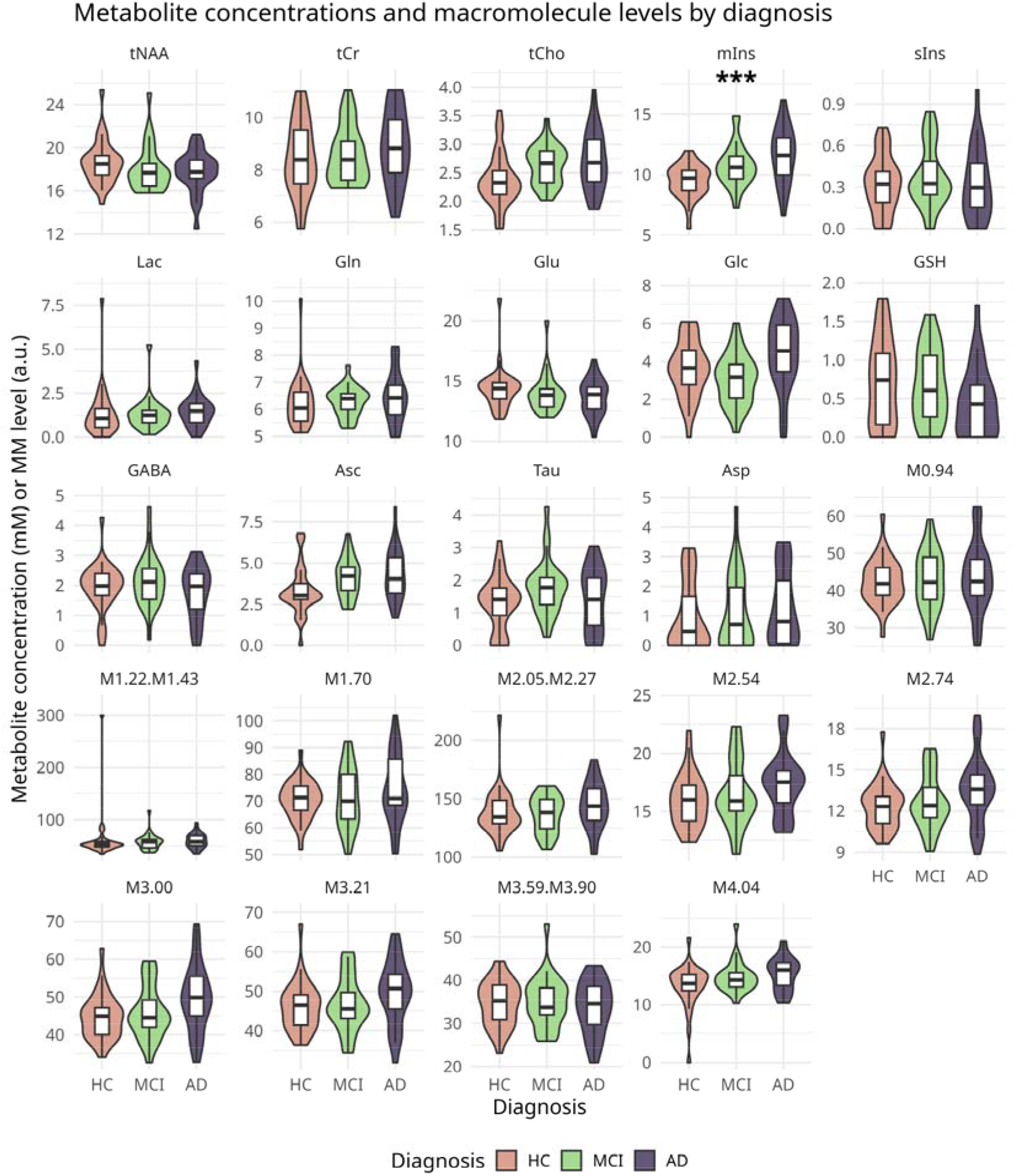
Metabolite concentrations and MM levels across diagnoses. Although many differences were not statistically significant after adjustment for age and multiple comparisons, descriptive trends were broadly consistent with previous reports. MRS quantification revealed higher levels of tCho, mIns, Asc, and reduced levels of Glu and GSH. Moreover, higher levels of M1. 70, M2. 54, M3. 00, M3. 21 and M4. 04 were found in AD. Note, that the AD participants are significantly older than the MCI or HC and the differences may be due to age. ^*^: p<0. 05, ^**^: p<0. 01, ^***^: p<0. 001, ^****^: p<0. 0001, n. s. non significant, based on adjusted p-values.

GSH and Tau were excluded from the classification models because of the missing values. Five missing values were imputed for sIns, two for GABA, Glc, and Lac, each, and one for Asc, M1.22, M3.90, and M4.04, each. The complete distribution of missing values before imputation is reported in Supporting Information Table S2.

### Classification Models and Metrics

The averaged training scores for the three pairwise OPLS-DA comparisons are shown in Supporting Information Figure S1. Classification performance on held-out MCCV test data is reported separately in Table 2, and additional OPLS-DA model diagnostics across MCCV rounds are shown in Supporting Information Figure S2.

**Table 2:** Classification metrics for each pairwise comparison. Values are reported as mean ± SD across 100 MCCV iterations unless otherwise indicated. R^2^X(cum), R^2^Y(cum), and Q^2^(cum) denote the cumulative values obtained from each fitted OPLS-DA model and are summarized across MCCV iterations.

|  | AD VS HC | MCI VS HC | AD VS MCI |
| --- | --- | --- | --- |
| **Q<sup>2</sup> (CUM)** | 0.31 ± 0.11 | -0.46 ± 0.25 | -0.23 ± 0.20 |
| **R<sup>2</sup>X (CUM)** | 0.70 ± 0.02 | 0.68 ± 0.02 | 0.71 ± 0.03 |
| **R<sup>2</sup>Y (CUM)** | 0.72 ± 0.03 | 0.40 ± 0.06 | 0.53 ± 0.05 |
| **F1 SCORE** | 0.66 ± 0.18 | 0.43 ± 0.16 | 0.61 ± 0.21 |
| **SENSITIVITY** | 0.76 ± 0.24 | 0.62 ± 0.26 | 0.60 ± 0.26 |
| **SPECIFICITY** | 0.79 ± 0.13 | 0.53 ± 0.14 | 0.68 ± 0.17 |
| **AUC** | 0.88 | 0.60 | 0.67 |
| **ACCURACY** | 0.78 ± 0.11 | 0.55 ± 0.11 | 0.64 ± 0.17 |

The area under the curve (AUC) values were 0.88 for AD vs. HC, indicating very good discrimination, while MCI vs. HC and AD vs. MCI showed lower AUC values of 0.60 and 0.67, respectively, reflecting challenges in distinguishing these groups. AUROCs, F1 score, sensitivity, specificity, and accuracy across MCCV iterations are reported in Table 2 and Figure 3A, while ROC curves are shown in Figure 3B.

**Figure 3:**
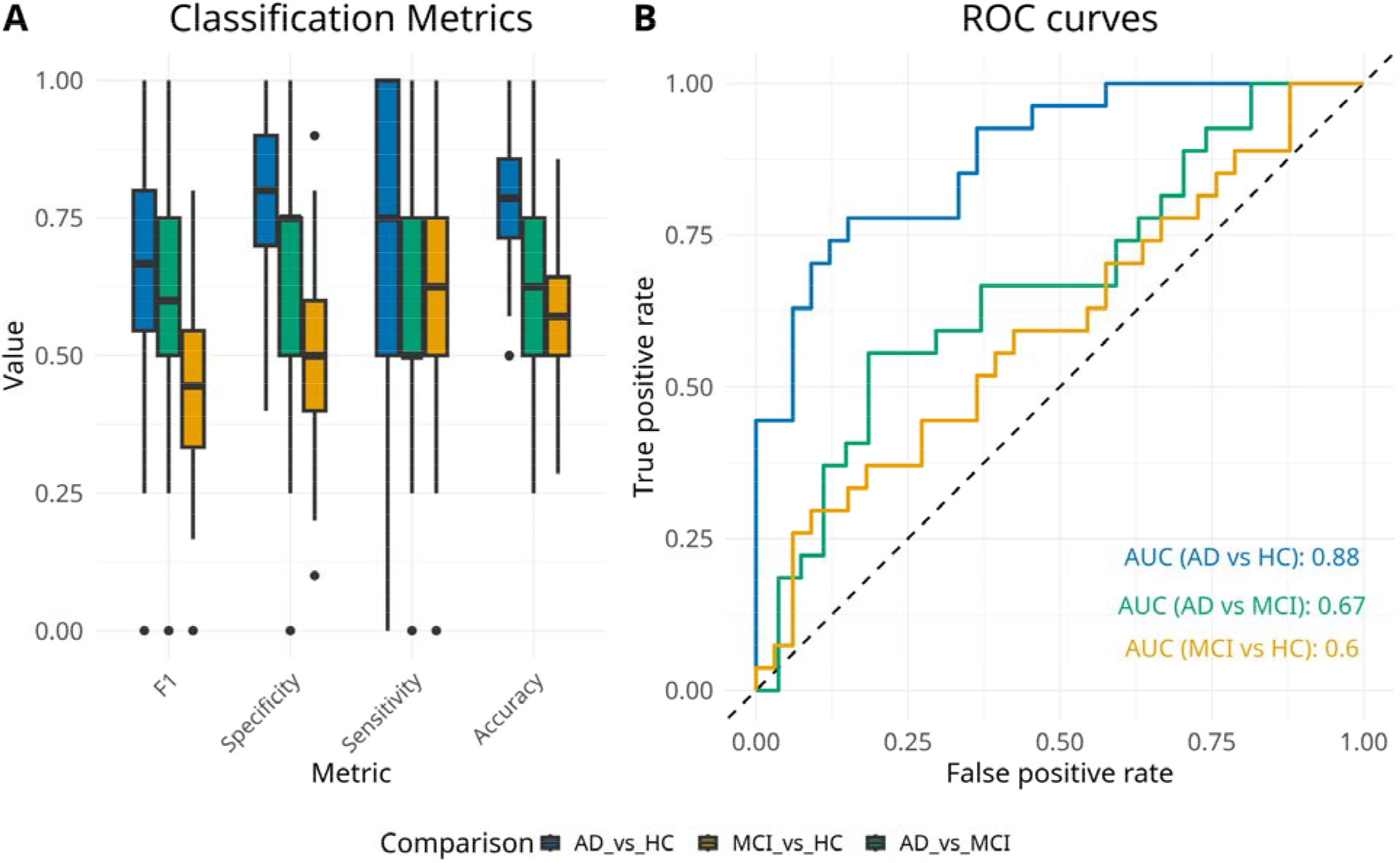
Classification performance of the pairwise OPLS-DA models. (A) Distribution of F1 score, specificity, sensitivity, and accuracy across MCCV iterations for AD vs HC, MCI vs HC, and AD vs MCI. (B) ROC curves for the three pairwise comparisons with the corresponding AUC values.

### Feature importance in the classification

The following features resulted in an aVIP higher than 1. For AD vs HC: Age, tCho, mIns, Asc, Glc, M2.54, M2.74, M3.00, M3.21, M4.04. For the classification AD vs MCI: Age, Glc, mIns, M3.00. For the classification MCI vs HC: Asc, Glc, tCho, mIns, M4.04. The aVIP values with the corresponding SD for all the metabolites, MMs, and for the three classifications are represented in Figure 4. The distribution of correct predictions across individuals and classifications is represented in Supporting Information Figure S3.

**Figure 4:**
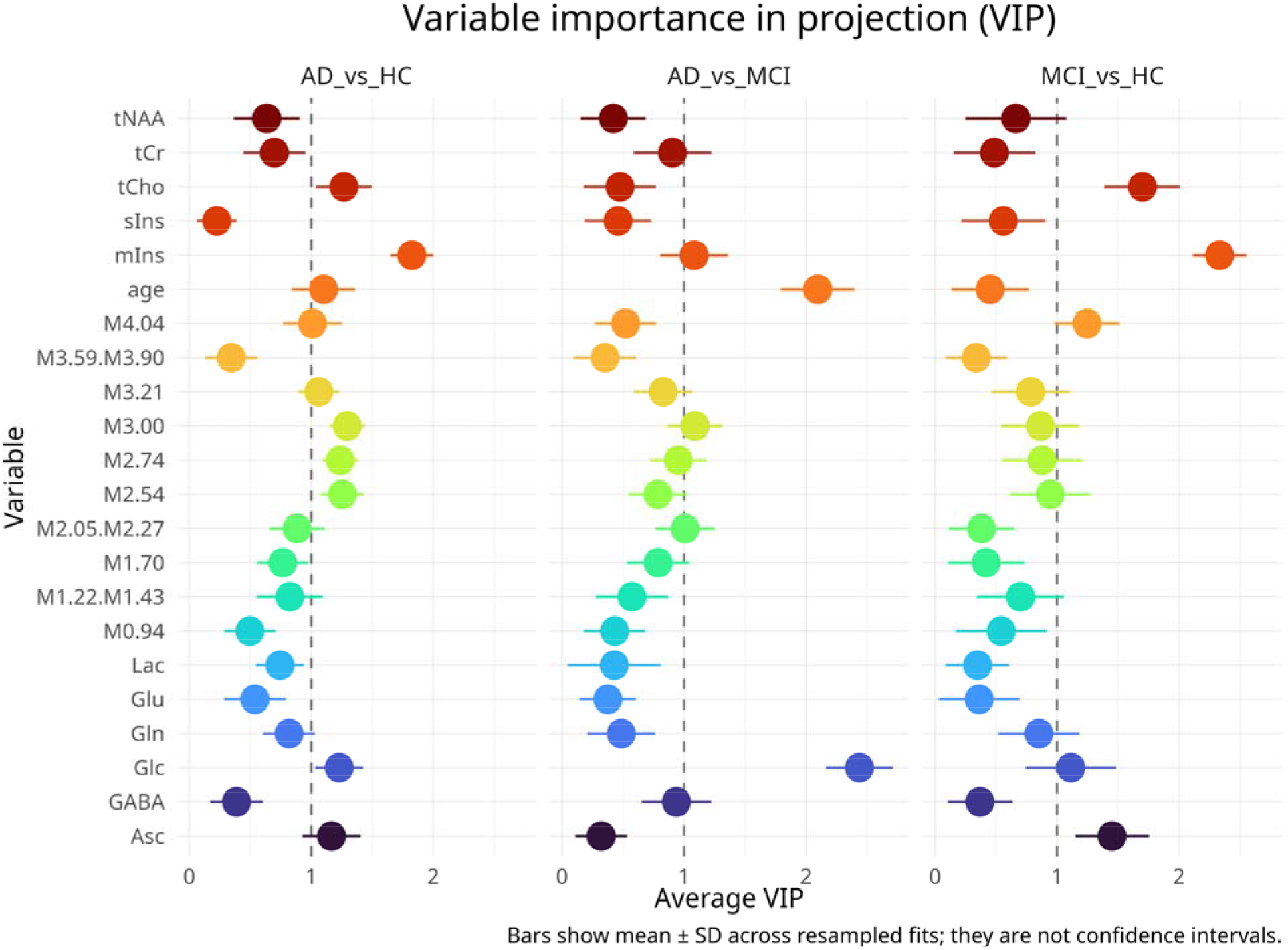
Variable Importance in Projection: The dots represent the average values from the MCCV rounds, while the line represents the standard deviation. Across the three comparisons, mIns was consistently identified as an important metabolite, while tCho showed high variable importance in the AD versus HC and MCI versus HC comparisons. Asc was similarly important for AD vs HC and MCI vs HC. M4. 04 and the MM peaks between 2. 54 ppm and 3. 21 ppm may have higher predictive potential than the others

## Discussion

In this study, we investigated the potential of using high-resolution 7 T ^1^H-MRS data to predict the diagnoses of MCI and AD in comparison with HC using a multivariate classification approach based solely on ^1^H-MRS-derived features and participant age. The model achieved robust performances in distinguishing AD from healthy controls, indicating that neurochemical alterations at later disease stages are sufficiently pronounced to be captured by this approach. In addition, the metabolites contributing most strongly to the classification were consistent with those reported in previous studies, providing insight into neurochemical alterations associated with the progression of dementia.

Classification performance was markedly lower for comparisons involving MCI. The combination of relatively high R^2^Y and low or negative Q^2^ suggests that MCI-related differences were captured in the training data but did not generalize, consistent with substantial neurochemical overlap with both HC and AD. This may reflect the etiological heterogeneity of MCI: some individuals represent prodromal AD, whereas others have different underlying causes, remain stable, or revert to normal cognition. A recent meta-analysis reported progression to dementia in 27% of population-based and 42% of clinical MCI cohorts over a mean follow-up of 5.2 years, while approximately 15% reverted to normal cognition[28]. These different underlying etiologies may therefore limit discrimination of MCI from both HC and AD.

The models’ aVIPs provide insight into diagnosis-related neurochemical profiles. However, the poor discrimination of MCI from AD or HC limits their clinical utility for early AD detection. Because diagnostic categories were based on neuropsychological assessment, and MCI is etiologically heterogeneous and may reflect causes other than AD pathology[29,30], longitudinal studies are needed to track neurochemical changes and distinguish markers of progression to AD from those associated with other MCI etiologies.

Studies performing similar classifications are rare. Munteanu et al. (2015) investigated the classification of AD and MCI versus controls using MRS data, tissue fractions, and volumes from the left and right hippocampi or parietal lobes. The models achieved AUC values of approximately 0.85 in several comparisons, with the highest performance obtained from models primarily based on tissue volumes after feature selection. Nevertheless, Munteanu et al. identified metabolites such as mIns and tCho as important discriminators, consistent with our findings. Additionally, some models identified tCr or tNAA as important features as well.

Marjańska et al.[8], investigated the neurochemistry of AD compared to HC and found, consistent with our study, that the most important metabolites distinguishing AD and HC neurochemistry were mIns, Asc, and tCho measured in the PCC. Moreover, the same study identified mIns and ascorbate as the most predictive metabolites for the classification of AD vs HC using a random forest classification.

Recently, two attention-based neural networks using the real spectrum achieved >90% accuracy for HC versus MCI or AD classification[31,32]. However, feature importance was inconsistent, limiting biological interpretability.

Although few metabolites remained significant after adjustment for age and multiple comparisons, the overall pattern was consistent with previous AD MRS findings: higher tCho, mIns and Asc and lower Glu and GSH[4,8,33], together with higher M1.70, M2.54, M3.00, M3.21 and M4.04. Because the AD group was older, some differences may be age-related. Age was therefore included explicitly in the OPLS-DA models. This is particularly relevant for MM resonances, which show age effects at 7 T, especially around 1.7 ppm in the PCC[34]. Accordingly, M1.70 was no longer significant after age adjustment and had low VIPs, suggesting that its apparent AD association may be age-related.

The aVIP analysis identified metabolites and MM peaks contributing to group discrimination (Figure 4), although only AD versus HC yielded significant classification and aVIPs from MCI-related comparisons should therefore be interpreted cautiously.

mIns, a marker linked to astrocytic activation and neuroinflammation[33], was the most predictive metabolite across comparisons and is consistently reported to be elevated in AD and MCI[5,33,35]. Clinically, mIns/Cr has been associated with MCI-to-AD conversion over 12 months (70% sensitivity, 90% specificity)[36]. However, increasing mIns across prodromal AD may make it more informative for progression than for cross-sectional discrimination. Longitudinal analysis of the same cohort showed increasing PCC mIns in MCI, with comparatively stable levels in HC and AD[37]. Etiological heterogeneity of clinically defined MCI may further obscure this relationship. Consistent with this, analyses of the same cohort showed better classification when mIns was combined with other metabolites than when used alone[17,38].

tCho, the second most important metabolite across classifications, is associated with membrane turnover, demyelination and possibly compensatory responses to acetylcholine deficits, and has repeatedly been linked to AD in the PCC[4,33].

Reduced tNAA levels have been reported in AD and MCI in previous studies. A recent review [4] of 85 publications investigating AD and MCI using ^1^H-MRS found that only one [39] study reported significantly lower levels of NAA in MCI and AD after absolute quantification and PVC. In most studies, tNAA ratios with metabolites such as mIns or tCr were used[4]. These metabolites have been reported to be increased in MCI and AD [4], thus the decrease of tNAA/tCr or tNAA/mIns ratios may be due to the sole increase of the concentration of the metabolite in the denominator. Our study supports this notion, as tNAA appears not to contribute significantly to the classification.

The identification of MM peaks among relevant features suggests that the MM background may contain disease-related information beyond metabolite concentrations. However, interpretation is challenging because MM resonances cannot generally be assigned uniquely to specific biochemical compounds[6,40] and overlap with several metabolite signals.

Several relevant MM peaks overlap with metabolite resonances, including Glu/Gln (M2.54, M2.74), tCr (M3.00), and tCho (M3.21). M4.04, which contributed to discrimination of HC from AD and MCI, also overlaps with mIns and Asc and lacks soft constraints in the PRaMM model. Although fit quality and CRLBs did not suggest major misattribution, some ambiguity in signal assignment remains. MM-related VIP findings should therefore be considered exploratory and require validation using dedicated MM acquisitions or improved MM modelling.

### Limitations and future development

Several limitations should be considered. First, the modest cohort size may reduce the stability of MCCV estimates, particularly for heterogeneous MCI cases, although balanced sampling ensured equal group representation within each pairwise classification. Second, the older age of the AD group may partly influence classification performance and VIP rankings. Age was included explicitly in the models, but residual confounding cannot be excluded.

Diagnosis was based primarily on neuropsychological assessment rather than biological markers. Biomarker-based definitions incorporating amyloid or tau may therefore better characterize AD-related neurochemical changes and help interpret consistently misclassified cases. Moreover, pairwise OPLS-DA treats HC, MCI, and AD comparisons independently and cannot model their potential ordering along a disease trajectory.

Diagnosis-stratified kNN imputation may also have reduced within-group variability or introduced diagnosis-dependent bias, although few values were affected. Finally, the cross-sectional design limits separation of inter-individual variability from longitudinal disease-related change. Longitudinal, biomarker-confirmed studies are therefore needed to validate neurochemical trajectories and assess whether OPLS-DA can support differential diagnosis between AD and clinically overlapping neurodegenerative disorders.

## Conclusion

In this study, we characterized for the first time individual MM components within the AD spectrum. We observed neurochemical patterns consistent with previously reported ^1^H-MRS biomarkers, including established metabolites such as mIns and tCho, as well as Asc, which has recently emerged as a potentially relevant marker. Using high-resolution 7 T ^1^H-MRS data and an OPLS-DA analysis, we achieved reliable classification performance in distinguishing AD from healthy controls and identified the metabolites and MM peaks showing the strongest contributions to AD-related neurochemical alterations. These findings demonstrate that ^1^H-MRS, when combined with advanced quantification and multivariate approaches such as OPLS-DA, can provide a comprehensive and non-invasive characterization of the biochemical changes associated with AD. Beyond its diagnostic potential, this approach contributes to a deeper understanding of the neurochemical processes underlying disease progression and highlights the value of integrating metabolic information into multimodal biomarker frameworks. As such, it represents an important step toward more precise characterization of the AD continuum and may contribute to future strategies for earlier detection, patient stratification, and monitoring of disease-related changes.

### Open Data Statement

MR data and concentrations will be made open source at the end of the (NeuroMET) study. Exemplificative model code is made available on GitHub (https://github.com/0rC0/AD_OPLSDA)

### Informed consent statement

All participants gave written informed consent before participating in the study.

## Statement on the use of generative AI

Generative AI tools were used exclusively to improve grammar, readability, and conciseness of the manuscript. No scientific content, data interpretation, or evaluative text was generated by AI. All analyses, results, and scientific conclusions were produced, reviewed, and verified by the authors, who take full responsibility for the accuracy and integrity of the manuscript.

