## Supporting Information for "Neurochemical fingerprinting of the Alzheimer’s disease continuum using 7 T short-echo-time brain single voxel ^1^H-MRS"

#### S1. MRS processing and quantification

We used a dataset of single-voxel short echo-time 7 T brain ^1^H-MRS data from the PCC. To utilize all information from the spectra for classification, we performed a simultaneous quantification of metabolites and MM using our previously proposed PRaMM model [1], which was demonstrated to obtain result accuracy comparable to those obtained with in-vivo measured MM components in the same voxel.

To achieve fully localized spectra, the raw MRS data underwent preprocessing using a custom MATLAB-based pipeline also used in previous studies [2–4]. The initial step involved averaging each odd acquisition with the subsequent even acquisition, resulting in a fully localized spectrum. Subsequently, we performed a weighted, phase-corrected coil combination, followed by frequency alignment to the NAA methyl peak at 1.99 ppm and averaging of the aligned signals.

We employed SPM12 [5] to perform a three-tissue segmentation of the MP2RAGE images. The MRS voxel masks were subsequently applied to the tissue masks using an in-house Python script. From the resulting MRS voxel tissue masks, we calculated the tissue fractions that were used for the partial volume correction (PVC).

The preprocessed spectra were analyzed and quantified using LCModel v6.3 [6]. We generated a simulated basis set using VeSPA v0.9.3 [7], which included the following metabolites: alanine (Ala), Asc, aspartate (Asp), phosphoryl-choline (PCho), creatine (Cr), phosphocreatine (PCr), γ-aminobutyric acid (GABA), glucose (Glc), glutamine (Gln), glutamate (Glu), glutathione (GSH), mIns, lactate (Lac), NAA, scyllo-inositol (sIns), taurine (Tau), N-acetylaspartylglutamic acid (NAAG) and glycerylphosphorylcholine (GPC). To correct for coil load, LCModel used a non-water-suppressed spectrum to scale the resulting peak areas, which was acquired in each volunteer with only four averages, but otherwise identical settings to the spectral acquisition.

Additionally, to model the MM components, we incorporated a model of simulated peaks and associated soft constraints (CHSIMU and CHRATIO, respectively) from our previous work [1].

We visually inspected the spectral quantifications and calculated the fit quality number (FQN, Wilson et al., 2019) to ensure spectral quality. Spectra with evident lipid contamination or artifact were excluded.

Next, we extracted metabolites and MM peak intensities from the LCModel COORD files. Metabolite or MM levels with Cramér–Rao lower bounds (CRLBs) of 999 were considered missing values and marked as NA. To obtain absolute metabolite concentrations, we corrected the LCModel output peak intensities for relaxation times and applied PVC using the CSF fraction from the three-tissue segmentation, following the experts’ consensus guidelines from Near et al. (2021). This process allowed us to determine the millimolar (mM) concentrations of metabolites.

Because each MM peak is composed of the resonance of multiple protons from the side chains of amino acids of the brain proteins, relaxation times for MM would be poorly defined; therefore, we chose to perform only water scaling and PVC to the MM values. However, the proposed classification model evaluates the variance of a variable across individuals and diagnoses, making it robust in handling heterogeneous data.

Table S1: MR acquisition parameters

|  | MRI |  | MRS |
| --- | --- | --- | --- |
| MR Scanner | Siemens MAGNETOM 7 T | | |
| RF coil | NOVA Medical 1TX/32RX head coil | | |
| Pulse Sequence | MP2RAGE | **Pulse Sequence** | SPECIAL |
| TR / TE (ms) | 5000 / 2.51 | **TR / TE (ms)** | 6500 / 9.0 |
| TI1 / TI2 (ms) | 900 / 2700 | **Number of acquisitions (NA)** | 64 |
| Bandwidth | 250 Hz/pixel | **VOI size** | 20 × 20 × 20 mm³ |
| Flip angles (α1 / α2) | 7°/5° | **Number of sample points** | 2048 |
| Isotropic resolution | 0.75 mm | **Acquisition duration** | 512 ms |
| Field of view (FoV) | 240 × 240 × 180 mm³ | **Excitation bandwidth** | 4000 Hz |
| GRAPPA acceleration factor | 2 | **Water suppression bandwidth** | 60 Hz |
| Number of reference lines | 32 | **Number of transients** | 64 |
| Total acquisition time | 11 minutes and 17 seconds | **Vector size** | 4096 |
|  |  | **Acquisition Bandwidth (Hz)** | 4000 |

### S2. Excluded Spectra

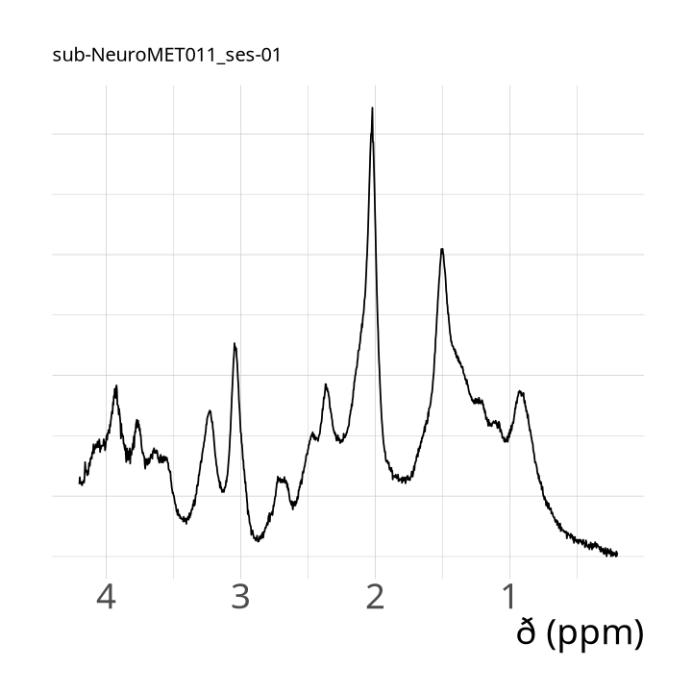

Reason of exclusion: major lipids contamination at about 1.2-1.5 ppm

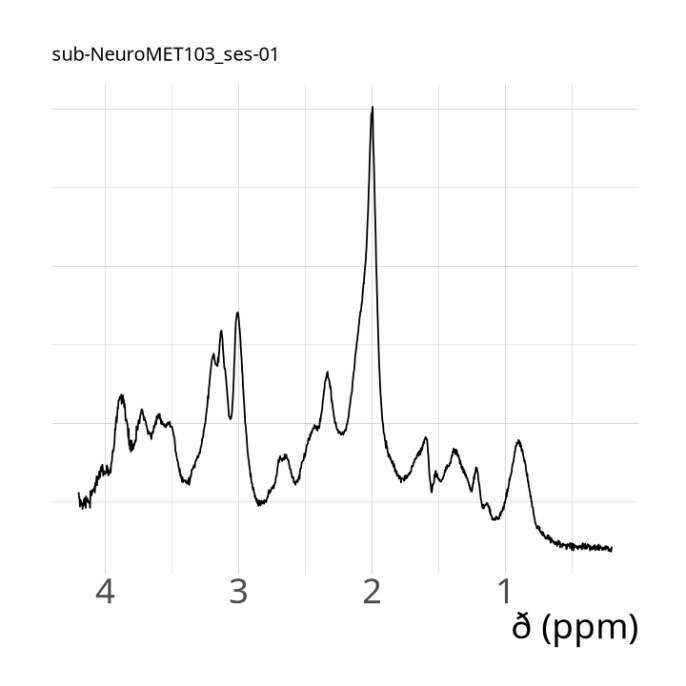

Reason of exclusion: major artifact at 3.2 ppm. It was visually verified that the second peak was not because a suboptimal coregistration during preprocessing.

#### S3. Missing values

Table S2: Missing values before imputation. Missing measurements for each metabolite and macromolecular (MM) component, reported overall and by diagnostic group. Variables with more than 10% missing values (GSH, Tau, and sIns) were excluded from classification analyses. Remaining missing values were imputed using diagnosis-stratified k-nearest neighbor (kNN) imputation (k = 5) including age as a predictor. Imputation was applied exclusively to OPLS-DA analyses.

| Metabolite/MM | Missing values |
| --- | --- |
| GSH | 15 |
| Tau | 8 |
| sIns | 5 |
| GABA | 2 |
| Glc | 2 |
| Lac | 2 |
| Asc | 1 |
| M1.22 | 1 |
| M3.90 | 1 |
| M4.04 | 1 |

### S4. Concentrations

Table S3: Metabolite concentrations, macromolecular peak intensities, and ANCOVA results across diagnostic groups. Metabolite concentrations are reported in millimolar units (mM), whereas macromolecular (MM) components are reported as water-scaled and partial-volume-corrected peak intensities. Values are presented as mean ± standard deviation for each diagnostic group. Group effects were assessed using analysis of covariance (ANCOVA) with age as covariate. Reported p-values were adjusted for multiple comparisons using false discovery rate (FDR) correction. AD: Alzheimer’s disease; MCI: mild cognitive impairment; HC: healthy controls; MM: macromolecules; ANCOVA: analysis of covariance; FDR: false discovery rate.

| Metabolite | Concentrations | |  | |  | | ANCOVA | |  |
| --- | --- | --- | --- | --- | --- | --- | --- | --- | --- |
|  | HC | | MCI | | AD | | F | | padj |
| tNAA | 18.63 ± 2.00 (n=33) | | 18.06 ± 2.14 (n=27) | | 17.77 ± 1.91 (n=27) | | 1.46 | | 0.510 |
| tCr | 8.45 ± 1.38 (n=33) | | 8.57 ± 1.08 (n=27) | | 8.86 ± 1.38 (n=27) | | 0.77 | | 0.699 |
| tCho | 2.38 ± 0.46 (n=33) | | 2.63 ± 0.34 (n=27) | | 2.71 ± 0.52 (n=27) | | 4.72 | | 0.092 |
| mIns | 9.47 ± 1.34 (n=33) | | 10.70 ± 1.72 (n=27) | | 11.61 ± 2.30 (n=27) | | 11.07 | | 0.001 |
| sIns | 0.35 ± 0.19 (n=31) | | 0.39 ± 0.21 (n=26) | | 0.37 ± 0.24 (n=24) | | 0.25 | | 0.894 |
| Lac | 1.40 ± 1.40 (n=32) | | 1.30 ± 0.95 (n=27) | | 1.51 ± 0.85 (n=26) | | 0.22 | | 0.894 |
| Gln | 6.23 ± 0.90 (n=33) | | 6.29 ± 0.54 (n=27) | | 6.46 ± 0.94 (n=27) | | 0.62 | | 0.740 |
| Glu | 14.35 ± 1.72 (n=33) | | 13.98 ± 1.62 (n=27) | | 13.65 ± 1.49 (n=27) | | 1.58 | | 0.508 |
| Glc | 3.70 ± 1.36 (n=32) | | 3.02 ± 1.31 (n=27) | | 4.61 ± 1.64 (n=26) | | 8.01 | | 0.008 |
| GSH | 0.85 ± 0.51 (n=28) | | 0.75 ± 0.43 (n=24) | | 0.63 ± 0.41 (n=19) | | 1.31 | | 0.510 |
| GABA | 2.00 ± 0.73 (n=32) | | 2.14 ± 0.87 (n=27) | | 1.84 ± 0.82 (n=26) | | 1.00 | | 0.597 |
| Asc | 3.45 ± 1.34 (n=32) | | 4.15 ± 1.19 (n=27) | | 4.31 ± 1.51 (n=27) | | 3.45 | | 0.175 |
| Tau | 1.56 ± 0.65 (n=28) | | 1.78 ± 0.85 (n=27) | | 1.64 ± 0.82 (n=23) | | 0.59 | | 0.740 |
| Asp | 1.59 ± 1.09 (n=20) | | 1.74 ± 1.19 (n=17) | | 1.73 ± 1.14 (n=20) | | 0.11 | | 0.894 |
| M0.94 | 42.50 ± 6.10 (n=33) | | 42.89 ± 7.77 (n=27) | | 43.54 ± 9.17 (n=27) | | 0.13 | | 0.894 |
| M1.22.M1.43 | 61.45 ± 44.32 (n=33) | | 57.25 ± 15.82 (n=27) | | 59.46 ± 13.71 (n=27) | | 0.15 | | 0.894 |
| M1.70 | 70.49 ± 7.08 (n=33) | | 71.08 ± 11.10 (n=27) | | 74.72 ± 12.70 (n=27) | | 1.38 | | 0.510 |
| M2.05.M2.27 | 138.12 ± 19.79 (n=33) | | 137.12 ± 15.64 (n=27) | | 143.96 ± 20.07 (n=27) | | 1.09 | | 0.584 |
| M2.54 | 15.89 ± 2.23 (n=33) | | 16.49 ± 2.88 (n=27) | | 17.37 ± 2.70 (n=27) | | 2.49 | | 0.267 |
| M2.74 | 12.33 ± 1.64 (n=33) | | 12.65 ± 2.00 (n=27) | | 13.56 ± 2.40 (n=27) | | 2.98 | | 0.225 |
| M3.00 | 44.66 ± 5.86 (n=33) | | 45.82 ± 6.98 (n=27) | | 49.62 ± 8.78 (n=27) | | 3.80 | | 0.158 |
| M3.21 | 46.01 ± 6.12 (n=33) | | 46.64 ± 6.83 (n=27) | | 49.58 ± 8.04 (n=27) | | 2.24 | | 0.302 |
| M3.59.M3.90 | 34.64 ± 5.16 (n=33) | | 35.01 ± 5.92 (n=27) | | 33.68 ± 6.06 (n=27) | | 0.42 | | 0.834 |
| M4.04 | 13.63 ± 2.94 (n=32) | | 14.78 ± 2.68 (n=27) | | 15.32 ± 2.85 (n=27) | | 2.77 | | 0.235 |

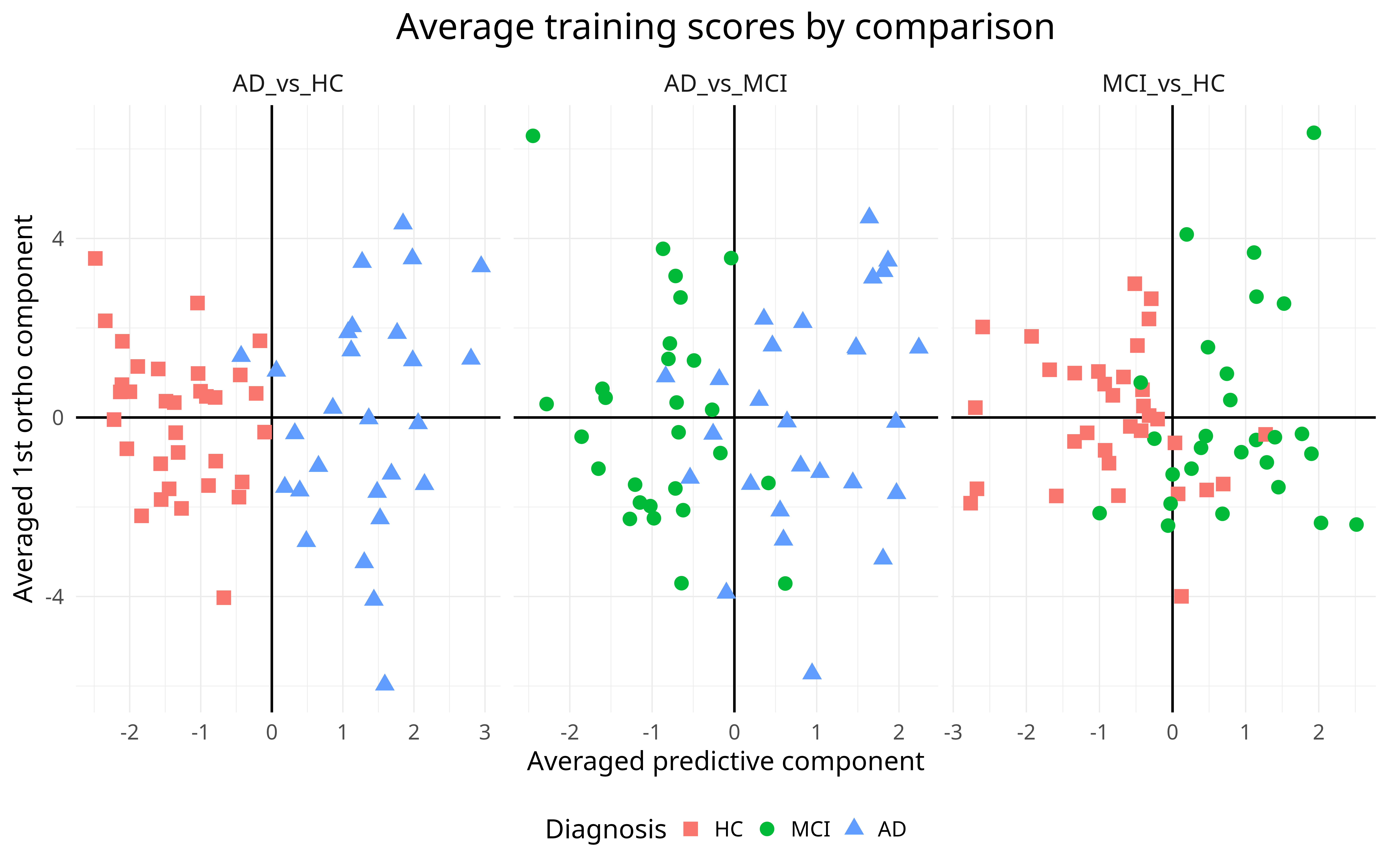

Figure S1: OPLS-DA Scores Plots. The predictive component is represented on the x-axis, while the first orthogonal component is depicted on the y-axis. Alzheimer's disease (AD) vs vs healthy controls (HC) shows the clearest visual separation, whereas greater overlap between diagnostic groups is observed for AD vs mild cognitive impairment (MCI) and MCI vs HC. OPLS-DA scores are derived from the training data and therefore illustrate the structure learned by the model rather than its performance on unseen data; visual group separation may consequently appear stronger than in test data.

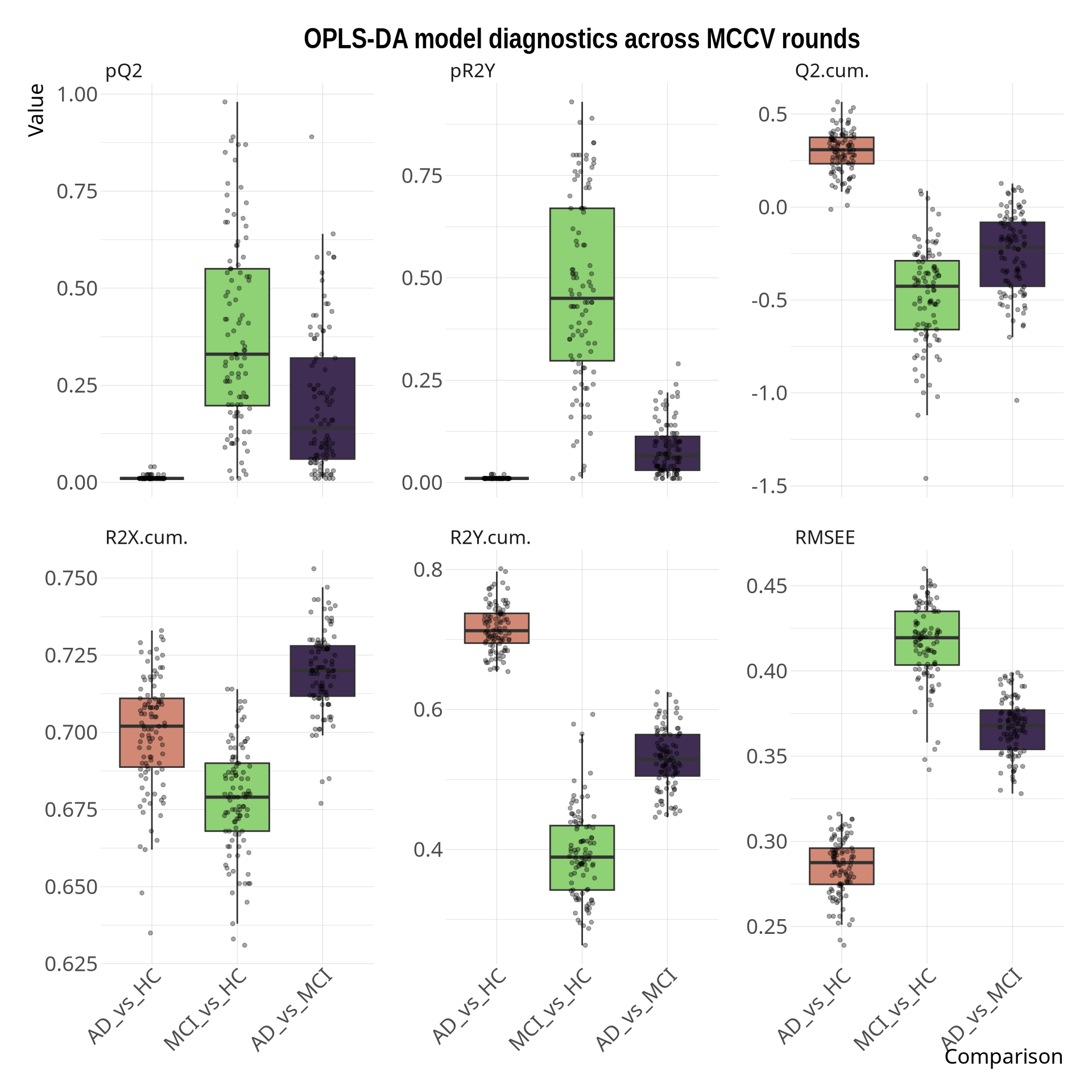

Figure S2: OPLS-DA model diagnostics across 100 MCCV rounds. The AD vs HC models demonstrated consistently positive Q2(cum) values and significant permutation-test results, indicating robust predictive performance. In contrast, models involving MCI showed markedly reduced predictive ability, with predominantly negative Q2(cum) values despite moderate R2Y(cum), suggesting that a substantial proportion of the explained variance was not generalizable to unseen data. This pattern is consistent with the greater neurochemical overlap between MCI and the other diagnostic groups.

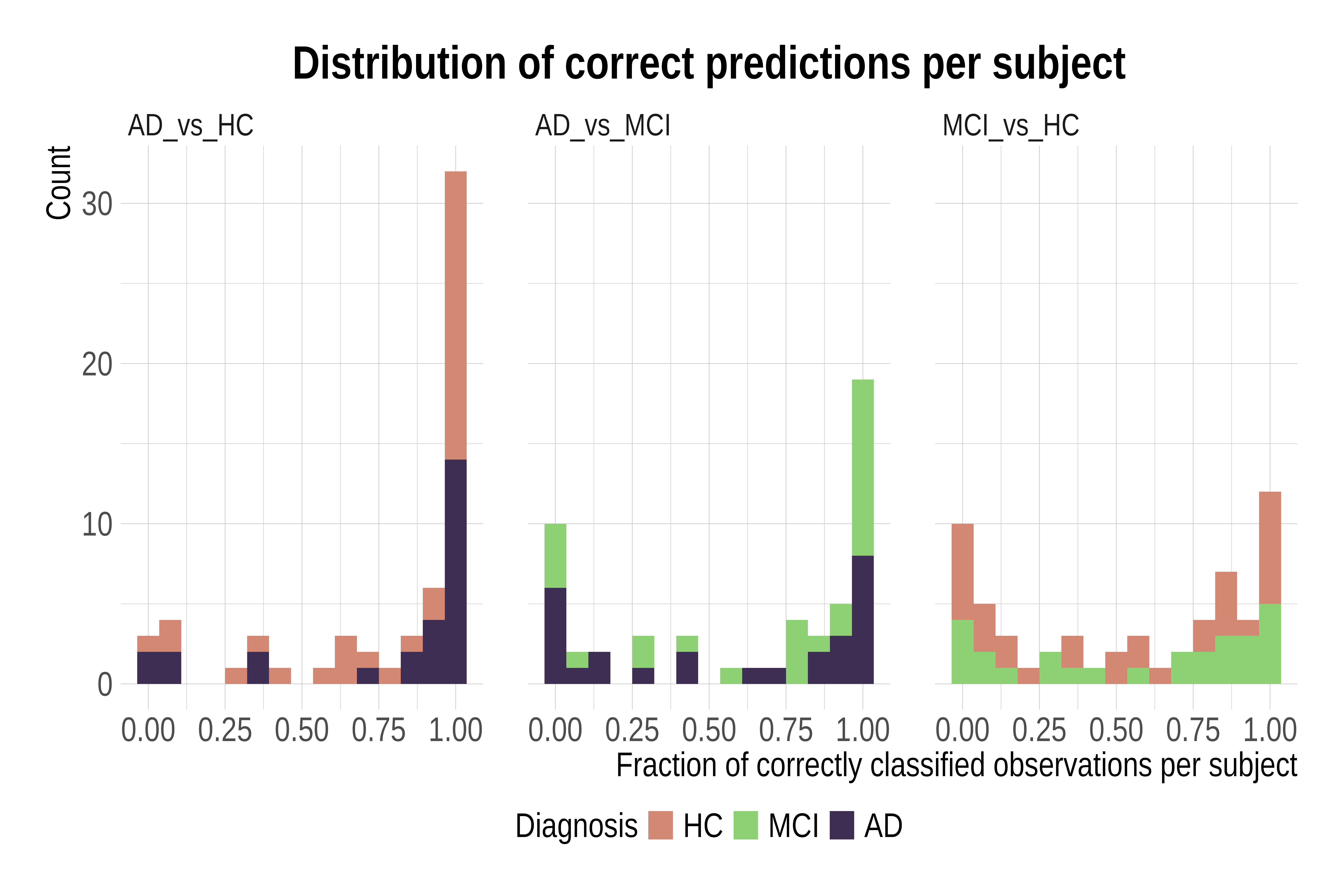

Figure S3:Fraction of correct classifications per subject across comparisons. For the AD vs HC comparison, most participants were consistently classified correctly, indicating robust separation between diagnostic groups. In contrast, higher variability in the AD vs MCI and MCI vs HC comparisons reflects overlapping neurochemical profiles and transitional characteristics of MCI along the Alzheimer’s disease continuum.
